# Immediate effects of suspension walker support on locomotor behavior of pre-walking infants with Down syndrome

**DOI:** 10.64898/2026.10.02.26364604

**Authors:** Christina M. Hospodar, Heather A. Feldner, Julia E. Looper, Kari S. Kretch

**Affiliations:** University of Southern California; University of Washington; University of Puget Sound

## Abstract

**Aims:** Our primary aim was to determine the immediate effects of using a supported stepping device on the real-time locomotor behavior of pre-walking infants with Down syndrome (DS). Our secondary aim was to characterize the role of the caregiver in facilitating infant locomotor behavior by quantifying infant-initiated and caregiver-initiated walker movement, and to test whether walker movement encourages stepping behavior.

**Methods:** Pre-walking infants with DS played with their caregiver in two conditions—with and without assistance from a wheeled suspension walker. To examine changes across development, we recorded infants across one to four sessions from 10 to 19 months of age.

**Results:** Across developmental stages, we found that suspension walkers robustly increased infants’ locomotor behavior: The walker increased upright time, percent time stepping, stepping rate, upright bout rate, percent time locomoting, and total bout rate. Most walker movement was initiated by caregivers, and infants stepped more while the walker was moving compared to stationary.

**Conclusions:** Even without practice, pre-walking infants with DS were able to take advantage of the suspension walker to increase their locomotor behavior. Caregivers facilitated infants’ activity by moving the walker. Future work will examine the potential of overground suspension walkers for home-based intervention.

## Locomotor delays in Down syndrome

Locomotion expands infants’ opportunities for play, learning, and participation in the home and community by increasing access to people, places, and things. Delays in locomotion—such as those experienced by children with Down syndrome (DS)—compromise infants’ ability to engage with the physical and social environment, which may have cascading effects on other areas of development such as cognition and language (Yamauchi et al., 2019). DS is a genetic condition associated with cognitive, language, and motor impairments, including delayed locomotor skills. For example, children with typical development begin walking at 12 months on average (WHO Multicentre Growth Reference Study Group, 2006). By contrast, children with DS begin walking at an average of 24 months and as late as 40 months, meaning walking can be delayed by upwards of two years (Winders et al., 2019). Thus, accelerating walking onset is a common intervention goal for children with DS. However, even the most effective interventions (e.g., treadmill training) accelerate walking onset by about four months at the maximum (D. A. Ulrich et al., 2008). Therefore, even with intervention, there is a large gap in experience between children with DS and their typically developing peers.

What experiences are typically developing toddlers acquiring that pre-walking children with DS are not? After the onset of walking, typically developing 12-month-olds spend a majority (around 70%) of play time upright—double the amount of same-aged pre-walkers (Franchak et al., 2018). Upright time is valuable because upright postures expand the visual field and free the hands, increasing visual and manual access to the environment (Franchak et al., 2018, 2011; Karasik et al., 2012; Kretch et al., 2014). Moreover, infants who can walk move a lot. In laboratory observations, walking infants spend about 50% of their time in motion, accumulating around 4000 steps and initiating 450 bouts per hour (Hoch et al., 2019). We know relatively little about locomotor behavior in infants with DS, but recent research suggests that 10-25-month-old infants with DS spend much less time moving than typically developing peers—less than 5% of the time moving compared to approximately 10-30% (Sutter et al., 2026).

## Facilitating on-time access to mobility for infants with DS

How can families and clinicians close the gap such that children with DS have access to similar experiences as their same-aged typically developing peers? The ON Time Mobility framework proposes that, rather than focusing exclusively on accelerating walking onset, clinicians and families should also seek to facilitate experiences that provide access to mobility in a developmentally appropriate time frame (Sabet et al., 2022). That is, clinicians and families should encourage the use of assistive devices at ages when walking would emerge in typical development—earlier than traditional introduction—so that children with motor delays can self-initiate movement before being able to walk independently.

Powered mobility devices (e.g., Permobil Explorer Mini, modified ride-on cars) provide one such option, and have been used successfully by young children with DS to move without adult assistance (Hospodar et al., 2021; Logan et al., 2019; Plummer et al., 2021; Sabet et al., 2026). Children demonstrated high enjoyment of powered mobility, and device use increased sibling socialization (Logan et al., 2019, 2014). Some powered mobility devices that can be used in an upright position even support motor development by encouraging weight-bearing and challenging postural control (Hospodar et al., 2021; Logan et al., 2019; Sabet et al., 2026).

Another option that both provides immediate access to mobility and supports developing skills are non-powered overground devices, like walkers and gait trainers. Compared to powered devices, overground devices allow for practice that is more task-specific for learning to walk because they provide support for upright stepping. Supported stepping devices like gait trainers are commonly used by clinicians to improve walking skill and participation in children with disabilities (Low et al., 2011; Paleg & Livingstone, 2015). Pre-walking infants with DS can take upright steps in a walker when the walker is pushed in controlled trials (Hospodar et al., 2026), but it is unclear how infants with DS respond to the walker in more naturalistic, play-based settings. We do not know how infants’ upright stepping with the walker compares to their locomotor behavior without the walker, how caregivers might interact with the walker and potentially facilitate movement, or how movement of the walker relates to infants’ stepping behaviors.

## Current study

We aimed to determine the immediate effects of using a supported stepping device on the real-time locomotor behavior of pre-walking infants with DS. Our primary aim was to compare infants’ locomotor behavior with and without a suspension walker, across developmental time and locomotor stages (pre-crawling, crawling, cruising). Although a walker *can* facilitate locomotor behavior, it need not necessarily make infants move more and could potentially even impede movement. Crawling and cruising status were of particular relevance: The walker may not provide benefit to pre-crawling or crawling toddlers if they are unable to take upright steps in it, and toddlers who can crawl or cruise may be able to move the same amount or more without the walker than they do with it.

Our secondary aim was to characterize the role of the caregiver in facilitating infant locomotor behavior by quantifying caregiver-initiated and infant-initiated walker movement. Because infant stepping in not inherently tied to walker movement (i.e., infants can step in place when the walker is stationary or take no steps when then the walker is moving), we also compared infant stepping when the walker was stationary versus moving to determine if walker movement encourages stepping behavior.

## Methods

### Participants

We observed 31 pre-walking infants (unable to take 10 unsupported steps) with DS from March 2024 to May 2026 (see Table 1 for demographic and medical information). Data collection occurred in a laboratory playroom at the University of Southern California (USC). An a priori power analysis suggested a sample size of 20 infants was sufficient to detect moderate statistical associations.

**Table 1.** Demographic and medical information. All responses were recorded at the infant’s first session.

|  | *n* | % |
| --- | --- | --- |
| Sex assigned at birth |  |  |
| Female | 17 | 54.8% |
| Male | 14 | 25.2% |
| Race |  |  |
| Black or African-American | 1 | 3.2% |
| White | 15 | 48.4% |
| More than one race | 3 | 9.7% |
| Unknown or not reported | 12 | 38.7 |
| Ethnicity |  |  |
| Hispanic or Latino | 27 | 87.1% |
| Not Hispanic or Latino | 4 | 12.9% |
| Most common language spoken in household |  |  |
| English | 16 | 51.6% |
| Spanish | 13 | 41.9% |
| Other | 2 | 6.5% |
| Highest educational level in household |  |  |
| Less than high school | 3 | 9.7% |
| High school degree | 13 | 41.9% |
| Associates/bachelors degree | 10 | 32.3% |
| Postgraduate degree | 5 | 16.1% |
| Household income |  |  |
| \$1 - \$30,000 | 6 | 19.4% |
| \$30,001 - \$60,000 | 6 | 19.4% |
| \$60,001 - \$100,000 | 6 | 19.4% |
| \$100,001 - \$200,000 | 5 | 16.1% |
| \$200,001+ | 4 | 12.9% |
| Unknown or not reported | 4 | 12.9% |
| Type of Down syndrome |  |  |
| Trisomy 21 | 28 | 90.3% |
| Mosaic | 1 | 3.2% |
| Unknown | 2 | 6.4% |
| Frequency of physical therapy |  |  |
| Does not receive physical therapy | 5 | 16.1% |
| 1-3 times per month | 3 | 9.7% |
| 4-7 times per month | 13 | 41.9% |
| 8 or more times per month | 10 | 32.3% |

Infants were eligible to participate if they had no additional medical or orthopedic conditions that limited standing or walking. We tested infants at four age points: 10, 13, 16, and/or 19 months (sessions were scheduled up to one month following the target age; e.g., between the 10-month and 11-month birthday for the 10-month session). Adjusted age was used to determine session windows for five infants born pre-term. We enrolled infants at the earliest possible session, and families participated in as many sessions as possible. Nine infants participated in one session, nine participated twice, ten participated in three sessions, and three participated in all four sessions, for a total of *N* = 69 sessions (15 10-month, 20 13-month, 16 16-month, 18 19-month). Infants completed both conditions in 55 of the 69 sessions (the no-walker condition was added after data collection began and was missing for the first 10 sessions; an additional no-walker condition and three walker conditions were not completed due to infant fussiness). One additional infant enrolled but was too fussy to complete the 10-month session and did not return for another visit. We excluded 13-month sessions from two included participants because the infants were too fussy to complete the sessions.

The procedure was approved by the USC IRB (UP-22-01092). Caregivers provided informed consent at the first session and were able to withdraw or skip future sessions at any time. Families received a $25 Amazon gift card per session and were provided with free parking or transportation reimbursement. We recruited participants by contacting parents of infants with DS in the target age window who had been seen at Children’s Hospital Los Angeles. Protocols, processed data, analysis code, and video data of each session (according to caregiver permission) are shared on Databrary at [URL-TO-BE-ADDED].

### Procedure

The free play observation (analyzed here) was part of a larger data collection comparing infant stepping behaviors across two tasks (stepping while supported over a treadmill versus stepping while pushed overground in the suspension walker; see Hospodar et al., 2026 for full details).

#### Free play observation

We observed infants playing with their caregivers in our laboratory playroom (5.3m x 4.7m), which was outfitted with foam blocks, wooden benches, and an assortment of toys (Figure 1A). Dyads played for approximately six minutes (*M* = 6.1 minutes, range: 5.0 – 7.3 minutes) without any device (*no-walker condition*) and for approximately six minutes (*M* = 5.8 minutes, range: 5.0 – 6.8 minutes) while the infant used a suspension walker (*walker condition*). The order of the free play conditions was counterbalanced, and each free play condition occurred immediately after the infant completed four 30-second trials stepping while supported on a treadmill (prior to the no-walker condition) or in the suspension walker (prior to the walker condition). In the no-walker condition, parents were instructed to play as they normally would, and in the walker condition, parents were told they were free to move the walker or not. In both conditions, parents were told they could go anywhere in the room and interact with any objects.

**Figure 1.**
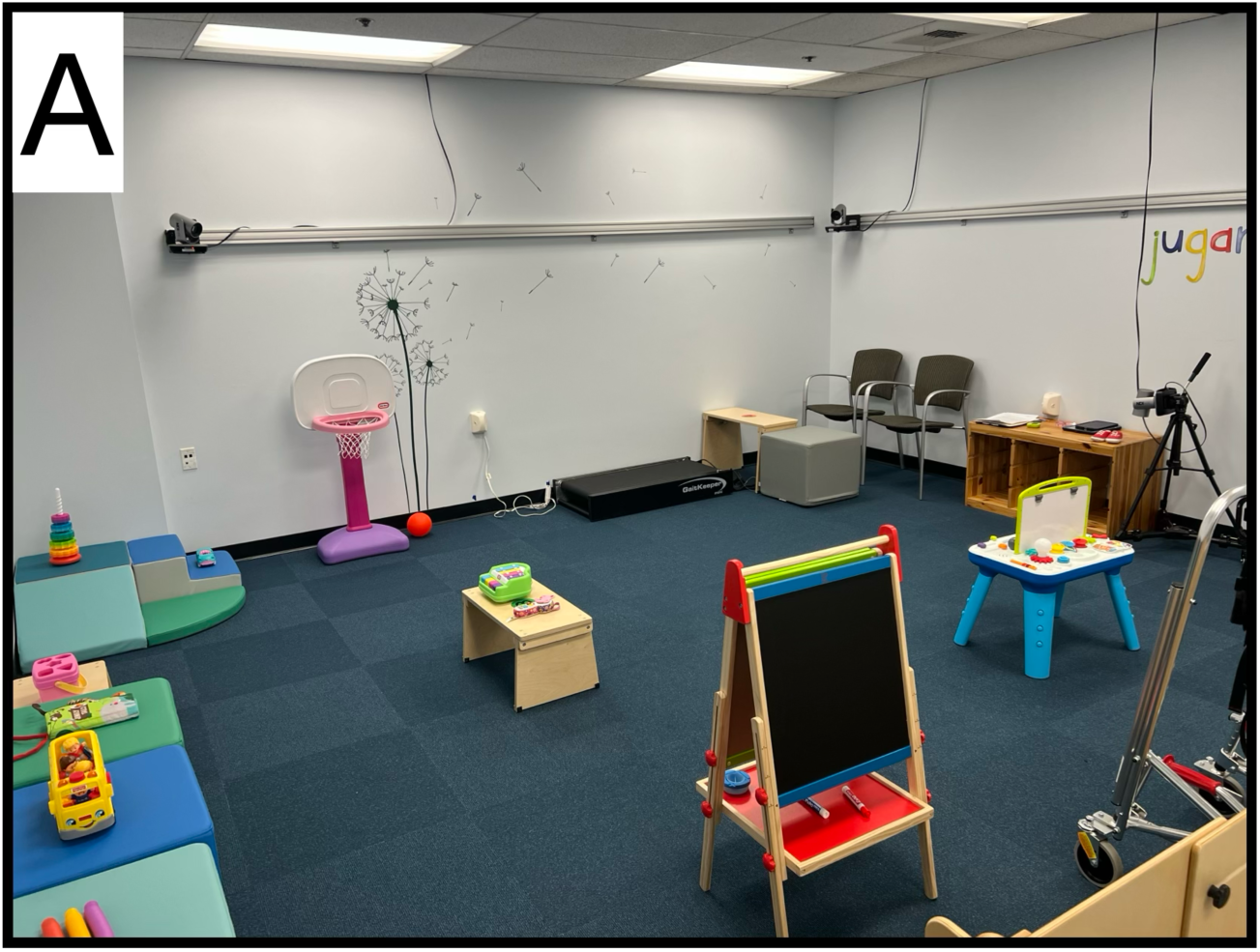
(A) Laboratory playroom and (B) infant in suspension walker. ***Figure 1b removed for preprint. Please contact the corresponding author for an image of infant in the suspension walker.***

In the walker condition, we used a Kaye suspension walker, a wheeled posterior walker with a mounted overhead bar (KAYE Products, Inc., 42” high, 24” x 23” footprint). The researcher outfitted the infant into an adjustable soft harness and then clipped the straps of the harness onto the overhead bar. The researcher adjusted the length of the harness straps so that the infant could maintain a fully upright position while bearing weight through the feet with knees and hips extended (Figure 1B). Infants wore lab-provided soft shoes with rubber soles during both free play conditions to avoid scraping the toes during the walker condition. All wheels of the walker were unlocked to facilitate movement in any direction.

At the end of the session, we verified infants’ crawling and cruising status. Infants were deemed “crawling” if they could move forward in a non-upright posture (e.g., hands-knees crawling, belly crawling, scooting, hitch crawling) for any distance. Infants were deemed “cruising” if they could cruise (take sideway steps holding on to furniture for support) at least five steps. We categorized infants into three locomotor status groups: *pre-crawlers* were not crawling or cruising (16 sessions), *crawlers* were crawling but not cruising (25 sessions), and *cruisers* were crawling and cruising (24 sessions); no infants could cruise but not crawl. Locomotor status is missing for four infants due to missing crawling status.

Free play was video recorded from four stationary cameras placed around the room, and an experimenter recorded infants with an iPad to obtain a close-up view of their movements. Videos were synced for annotation.

### Data coding

Videos were scored by trained coders in Datavyu video-coding software (https://datavyu.org). Some codes were only relevant to one condition, and some codes were coded in both conditions.

#### No-walker condition

Coders annotated infant *posture* frame-by-frame as upright (both feet on the support surface, with trunk angle greater than parallel to the surface) or non-upright (kneeling, prone, sitting, laying, or held). During non-upright time, coders annotated *non-upright locomotor bouts* by identifying periods of non-upright locomotion followed by at least a 1-s pause. Non-upright locomotion included hands-knees crawling, belly crawling, scooting, knee walking, and hitch crawling. To avoid counting postural shifts as locomotion, we only coded bouts that contained at least two distinct repetitions (e.g., two knee steps in hands-knees crawling, two lifts of the bottom for scooting). Note that these outcomes were not scored for the walker condition because infants were always upright in the walker and not able to engage in non-upright locomotion.

#### Walker condition

In the walker condition, coders annotated *walker movement* by identifying periods of movement across the floor followed by at least a 1-s pause. For each walker movement bout, coders identified the *movement source* as infant (self-propulsion) or caregiver (caregiver contacted the walker, pulled the infant’s hand(s)/body, or pulled the infant through a shared toy).

#### Both conditions

In both the walker and no-walker condition, coders annotated *upright locomotor bouts* by identifying periods of stepping followed by at least a 1-s pause. Although infants were all pre-walking, they could still take upright steps in the no-walker condition by cruising or stepping with handheld assistance. Bouts could contain any number of steps, including one. Bouts started the moment the foot lifted off the ground for the first step and ended when the foot regained contact with the ground for the last step. Coders also identified the *number of steps* in each upright bout. In the walker condition, we excluded steps if the infant lifted their foot for two seconds or longer; anecdotally, this behavior was common only in the walker condition due to the postural support provided by the walker so we chose to exclude these long steps to avoid artificially inflating the percent time spent stepping in the walker condition.

#### Reliability

For each measure, a primary coder annotated the entirety of the condition and a second coder independently annotated 20% of the condition. Inter-rater reliability was high for the total time upright (ICC= 1.00), the total time for non-upright locomotor bouts (ICC = 0.97), the total time for infant-initiated walker movement (ICC = 0.95), the total time for caregiver-initiated walker movement (ICC = 0.90), the total number of upright steps in the no-walker condition (ICC = 0.99), and the total number of upright steps in the walker condition (ICC = 0.92). Disagreements were resolved by discussion.

### Outcome measures

We calculated several outcome measures from the video-coded data. All outcome measures were normalized to task time to account for small differences in task time between infants and across conditions. For both conditions, *percent time stepping* was the sum of all upright locomotor bouts divided by the task time, and *percent time locomoting* was the sum of all locomotor bout durations (upright and non-upright locomotor bouts) divided by the task time. Note that these values were the same for the walker condition because all locomotion was upright. We also calculated the *stepping rate* (number of upright steps/task time), the *upright bout rate* (number of upright bouts/task time), and the *overall bout rate* (number of upright and non-upright bouts/task time). For the no-walker condition, *percent time upright* was the upright time divided by the task time (for the walker condition, this value was 100% by default).

For the walker condition, *percent time moving* was the total walker movement time divided by the task time. We also calculated the percent time moving separately for *infant-initiated* (total infant-initiated walker movement time divided by the task time) and caregiver-initiated (total caregiver-initiated walker movement time divided by the task time) movement. Finally, we determined *walker-dependent stepping rates* by calculating separate stepping rates for when the walker was stationary, moved by the infant, and moved by the caregiver (e.g., total steps taken while the walker was stationary divided by the total time the walker was stationary); if infants did not have at least 1-s total of any category of movement, we removed their data for that category because calculating a stepping rate for such a short period of movement would be unreliable.

### Data analysis

We ran separate statistical models for all outcome variables. For each outcome, we fit a linear mixed model with subject-level data using the *lme4* package in R. To account for individual differences, we included a random intercept for participant in all models. For significant interactions, we used the *emmeans* package to test follow-up comparisons.

For percent time upright and the six locomotor outcomes (percent time stepping, percent time locomoting, stepping rate, upright bout rate, overall bout rate), fixed effects included mean-centered infant age in months and condition (no-walker = 0, walker = 1), as well as the age×condition interaction. For percent time upright, infants were upright 100% of the time in the walker condition, but we still tested condition as a fixed effect to quantify the difference between conditions. We ran an additional set of models to test for order effects on the locomotor outcomes by adding in order and the order×condition interaction as predictors. To determine the effect of locomotor status, we re-ran the models for all six locomotor outcomes adding in a three-factor locomotor status variable (pre-crawlers, crawlers, cruisers, with pre-crawlers coded as the reference group) and the interactions between locomotor status and condition.

For percent time moving in the walker, fixed effects included mean-centered infant age in months, source of the movement (infant-initiated = 0, caregiver-initiated = 1), and the age×source interaction. For the walker-dependent stepping rate model, fixed effects included mean-centered infant age in months, movement category as a factor (stationary, infant-initiated movement, caregiver-initiated movement, with stationary coded as the reference group), and the interactions between age and movement category.

## Results

Our primary aim was to compare infants’ locomotor behavior between the two conditions and across age and locomotor status. Our secondary aim was to characterize the contribution of infants and their caregivers in producing walker movement and stepping behavior. We found no evidence that the effect of the walker depended on which condition was completed first (order×condition interaction *p*s > .54), so we ignored condition effects in following analyses.

### Comparing locomotor behavior across conditions, age, and locomotor status

#### Walker provided a large boost in upright time

At the average age, in the no-walker condition, infants spent 23% of the session upright, with considerable variability both across and within infants (Supplemental Figure 1A, intercept from Table 2A). Infants spent significantly more time upright with age, but the effect was small: With each additional month of age, infants spent an additional 3% of the time upright. Conversely, the condition effect was large: At the average age, the walker resulted in an additional 76% upright time.

**Table 2.** Summary of model results for postural and locomotor outcome variables. Values listed are coefficient estimates with the upper and lower bounds of the 95% confidence interval in parentheses. Age (months) was centered at the mean. Condition was dummy-coded (no-walker = 0, walker = 1). Thus, the intercept represents the average value for a child of the average age in the no-walker condition for models in section A. For models in section B, pre-crawling was coded as the reference condition and condition was dummy-coded (no-walker = 0, walker = 1). Crawling and cruising were coded as factors. Thus, the intercept represents the average value for a pre-crawler of average age in the no-walker condition. A random intercept for participant was included in all models. Cells are highlighted in green if significant. Asterisks denote *p*-values: \**p* ≤. .05, \*\**p* < .01, \*\*\**p* < .001.

|  | % Time Upright | % Time Stepping | Stepping rate (steps/min) | Upright bout rate (upright bouts/min) | % Time Locomoting | Overall bout rate (total bouts/min) |
| --- | --- | --- | --- | --- | --- | --- |
| **A. Age models** |  |  |  |  |  |  |
| Intercept | 23.32***<br>[17.49, 29.15] | 1.87*<br>[0.14, 3.60] | 1.85*<br>[0.11, 3.60] | 1.04***<br>[0.41, 1.66] | 5.43***<br>[3.46, 7.40] | 1.61***<br>[0.98, 2.24] |
| Age | 2.79***<br>[1.17, 4.41] | 0.42<br>[-0.10, 0.94] | 0.45<br>[-0.08, 0.98] | 0.19*<br>[0.01, 0.37] | 0.88**<br>[0.28, 1.49] | 0.27**<br>[0.08, 0.45] |
| Condition | 76.29***<br>[69.54, 83.04] | 10.37***<br>[8.14, 12.60] | 11.06***<br>[8.79, 13.34] | 4.71***<br>[3.95, 5.47] | 6.78***<br>[4.16, 9.40] | 4.13***<br>[3.35, 4.91] |
| Age×Condition | -2.85**<br>[-4.96, -0.73] | -0.16<br>[-0.86, 0.53] | -0.32<br>[-1.03, 0.39] | -0.10<br>[-0.34, 0.14] | -0.62<br>[-1.44, 0.20] | -0.18<br>[-0.43, 0.06] |
| **B. Locomotor status models** |  |  |  |  |  |  |
| Intercept | 11.48*<br>[0.37, 22.59] | 0.42<br>[-3.05, 3.89] | 0.34<br>[-3.13, 3.81] | 0.31<br>[-0.92, 1.54] | 0.81<br>[-3.10, 4.72] | 0.50<br>[-0.72, 1.72] |
| Age | 0.65<br>[-1.48, 2.78] | 0.09<br>[-0.59, 0.77] | 0.09<br>[-0.59, 0.77] | 0.03<br>[-0.20, 0.27] | 0.15<br>[-0.62, 0.91] | 0.05<br>[-0.18, 0.29] |
| Condition | 89.14***<br>[74.44, 103.84] | 13.91***<br>[8.98, 18.83] | 16.09***<br>[11.12, 21.05] | 6.27***<br>[4.60, 7.94] | 13.55***<br>[ 7.94, 19.17] | 6.08***<br>[4.40, 7.76] |
| Age×Condition | -0.39<br>[-3.20, 2.43] | 0.16<br>[-0.78, 1.10] | 0.06<br>[-0.89, 1.01] | 0.07<br>[-0.25, 0.39] | 0.10<br>[-0.97, 1.17] | 0.05<br>[-0.28, 0.37] |
| Crawling | 7.64<br>[-25.64, 10.49] | 0.23<br>[-4.19, 4.64] | 0.24<br>[-4.18, 4.66] | 0.21<br>[-1.33, 1.75] | 3.50<br>[-1.49, 8.49] | 0.52<br>[-1.02, 2.06] |
| Cruising | 25.46**<br>[ 9.11, 41.82] | 3.65<br>[-1.49, 8.80] | 3.86<br>[-1.28, 9.00] | 1.74<br>[-0.07, 3.55] | 9.28**<br>[3.48, 15.07] | 2.50**<br>[0.70, 4.31] |
| Crawl×Condition | -7.58<br>[-25.44, 10.28] | -3.02<br>[-9.08, 3.03] | -5.05<br>[-11.16, 1.06] | -1.31<br>[-3.36, 0.73] | -6.36<br>[-13.27, 0.54] | -1.64<br>[-3.71, 0.43] |
| Cruise×Condition | -28.08*<br>[-49.67, -6.48] | -5.50<br>[-12.73, 1.72] | -7.30*<br>[-14.59, -0.01] | -2.61*<br>[-5.06, -0.16] | -11.12**<br>[-19.36, -2.89] | -3.37**<br>[-5.84, -0.91] |

Upon adding in locomotor status and its interaction with condition as predictors, age was no longer significantly associated with upright time. There was a significant cruise×condition interaction. In the no-walker condition, cruisers spent more time upright than both pre-crawlers (difference = 25.5 percentage points, *p* = .008) and crawlers (difference = 17.8 percentage points, *p* = .046). Infants in all three groups were upright significantly more in the walker condition compared to no-walker condition (all *p*s < .001). However, the walker effect was smaller for cruisers compared to pre-crawlers (difference = 28.1 percentage points, *p* = .01) and crawlers (difference = 20.5 percentage points, *p* = .04; Supplemental Figure 1B) because cruisers spent more time upright in the no-walker condition.

#### Walker promoted locomotor activity across ages

There was a main effect of age on upright bout rate, percent time locomoting, and overall bout rate, suggesting older infants moved more overall and initiated more bouts (Table 2A and left panel of Figure 2). However, there was only a marginal effect of age on percent time stepping (*p*=.056) and no effect on stepping rate.

**Figure 2.**
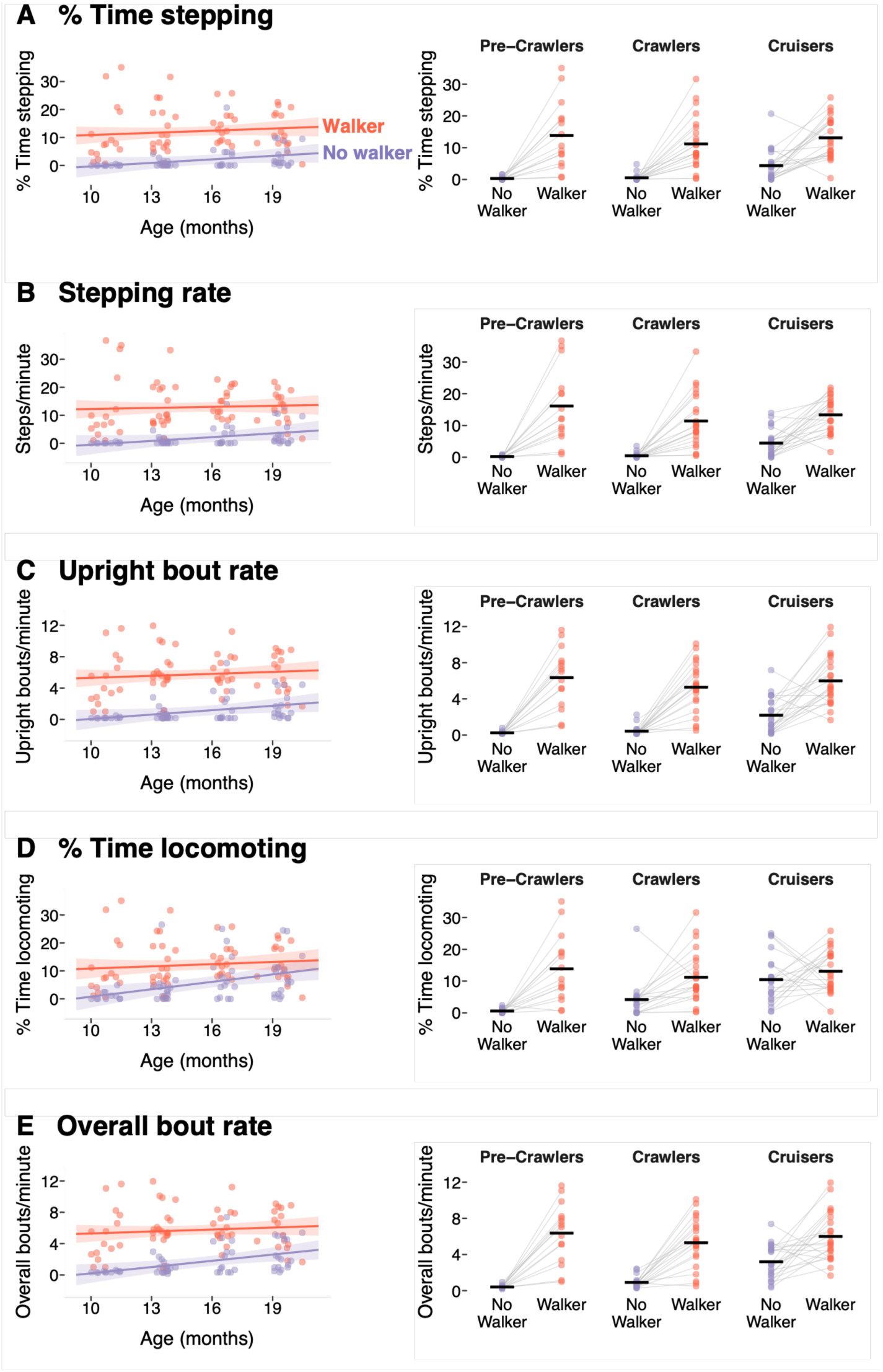
Effects of condition, age, and locomotor status on locomotor behavior. Left panel: Locomotor behavior as a function of age (x-axis) and condition (colors; purple = no-walker, red = walker). Banded lines represent the model estimation with 95% confidence interval. Circular points represent individual observations across visits from each infant. Right panel: Locomotor behavior across conditions for each locomotor status group. Circular points represent individual observations across visits from each infant. Gray lines connect data from the same infant across conditions. Horizontal black lines denote the mean for each condition per locomotor status.

At the average age, in the no-walker condition, infants moved very little. Infants spent only 2% of their time stepping, took 2 steps per minute, and initiated 1 upright bout per minute. When including their non-upright locomotion, infants in the no-walker condition spent 5% of their time locomoting and initiated 2 bouts per minute.

Across outcome measures, infants moved more in the walker condition compared to the no-walker condition: They spent more time stepping (an additional 10 percentage points), took more steps per minute (an additional 11 steps), and initiated more upright bouts per minute (an additional 5 bouts). In fact, 9/11 (82%) of the 10-month-olds, 7/16 (44%) of the 13-month-olds, 5/14 (36%) of the 16-month-olds, and 3/17 (18%) of the 19-month-olds took zero steps in the no-walker condition, but in every session, infants took at least three steps in the walker condition. Even when accounting for infants’ non-upright locomotion in the no-walker condition, infants still spent more time locomoting (an additional 7 percentage points) and initiated more locomotor bouts per minute (an additional 4 bouts) in the walker.

Across outcome measures, there was no age×condition interaction, suggesting the effect of the walker was not dependent on age.

#### Walker promoted locomotor activity regardless of locomotor status

When we added locomotor status and its interaction with condition as predictors, age was no longer a significant predictor of any outcome variable, and there was a significant cruise×condition interaction for stepping rate, upright bout rate, percent time locomoting, and overall bout rate. Follow-up comparisons indicated that for stepping rate, upright bout rate, and overall bout rate, infants moved significantly more in the walker condition compared to the no-walker condition regardless of locomotor status (all *p*s < .001); for percent time locomoting, pre-crawlers (*p* < .001) and crawlers (*p* = .003) moved significantly more in the walker condition, but cruisers did not (*p* = .33). For all four outcome variables, the walker effect was smaller for cruisers compared to pre-crawlers (all *p*s ≤.05).

### Characterizing the use of the suspension walker by infants and their caregivers

At the average age, infant-initiated walker movement accounted for only 3% of the total session time (Figure 3A, Table 3A). Caregiver-initiated walker movement accounted for significantly more walker movement than infant-initiated movement—an additional 10 percentage points. There was no effect of age on walker movement, or age×source interaction, suggesting total amounts of infant- and caregiver-initiated movement did not differ significantly across age. Although infant-initiated movement did not occur often, the majority of infants *could* move the walker on their own: 8/15 (53%) of the 10-month-olds, 16/20 (80%) of the 13-month-olds, 14/15 (94%) of the 16-month-olds, and 15/16 (94%) of the 19-month-olds had at least one bout of infant-initiated movement. In five of the sessions, caregivers never moved the walker and infants had no walker movement at all in only two of the sessions.

**Figure 3.**
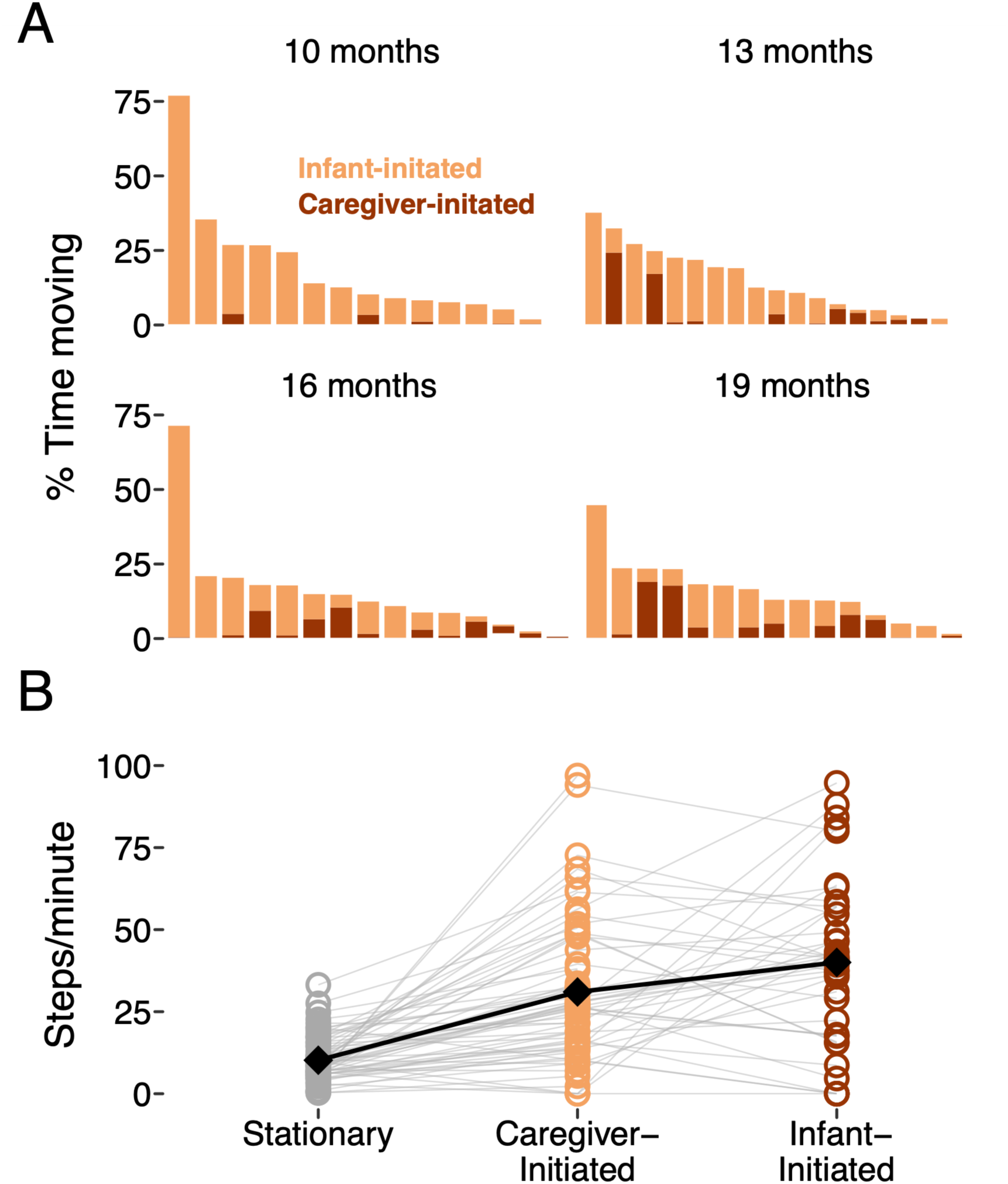
Walker movement and walker-dependent stepping rates. (A) Walker movement by age group. Each bar represents one session from one infant. Bars are ordered based on the total percent time moving within each age group. Colors within each bar denote infant-initiated movement (red) versus caregiver-initiated movement (yellow). (B) Walker-dependent stepping rates. Circular points represent individual observations across visits from each infant. Gray lines connect observations from the same session for the same infant. Black diamonds represent the mean stepping rate for each category.

**Table 3.** Summary of results for walker movement models. Age (months) was centered at the mean across models. For model A, infant-initiated movement was coded as the reference condition, and caregiver-initiated movement was coded as a factor. Thus, the intercept represents the infant-initiated percent time moving for a child of the average age. For model B, stationary was coded as the reference condition and infant-initiated and caregiver-initiated movement were coded as factors. Thus, the intercept represents the step rate while the walker was stationary for a child of the average age. A random intercept for participant was included in all models. Cells are highlighted in green if significant. Asterisks denote *p*-values: \**p* < .05, \*\**p* < .01, \*\*\**p* < .001.

|  | Estimate | 95% Confidence interval |
| --- | --- | --- |
| **A. Percent time moving** |  |  |
| Intercept | 2.94* | [0.20, 5.69] |
| Age | 0.37 | [-0.46, 1.20] |
| Caregiver-initiated | 9.54*** | [5.84, 13.23] |
| Age×Caregiver-initiated | -0.89 | [-2.05, 0.26] |
| **B. Walker-dependent stepping rates** |  |  |
| Intercept | 10.16*** | [ 5.74, 14.59] |
| Age | -0.07 | [-1.42, 1.28] |
| Infant-initiated | 28.47*** | [21.53, 35.41] |
| Caregiver-initiated | 21.14*** | [14.94, 27.35] |
| Age×Infant-initiated | 2.53* | [0.33, 4.72] |
| Age×Caregiver-initiated | 2.03* | [0.10, 3.96] |

At the average age, while the walker was stationary, infants took 10 steps per minute (Figure 3B, Table 3B). While the walker was moving, infants stepped much more: Infants took an additional 29 steps per minute when the walker was moved by the infant, and an additional 20 steps per minute when the walker was moved by the caregiver. Stepping rates were significantly higher when the walker was moving compared to stationary (*p*s < .001), but there was no significant difference between infant-initiated and caregiver-initiated walker movement (*p* = .11). During periods where the walker was stationary, stepping rates did not change with age, but stepping rates increased with age for both infant-initiated (an additional 2.5 steps per minute per month) and caregiver-initiated (an additional 2 steps per minute per month) movement periods.

## Discussion

We found that suspension walkers robustly increased the immediate locomotor behavior of pre-walking infants with DS: The walker increased upright time, percent time stepping, stepping rate, upright bout rate, percent time locomoting, and overall bout rate. Infants stepped more when the walker was moving, and the caregiver was the primary source of walker movement. Thus, although infants initiated their own stepping movements, caregivers played an active role in supporting their infants’ locomotor behavior via their engagement with the walker.

### The potential of an overground suspension walker to support everyday activity

In our prior work, pre-walking infants with DS demonstrated readiness for overground suspension walkers as early as 10 months, which aligns with prior work on treadmill stepping (B. D. Ulrich et al., 1992). The current study provides additional insight that infants of all locomotor statuses—including infants who could already step on their own (i.e., cruisers)—benefited from using the walker. This is notable because cruisers had multiple options for moving (crawling and cruising) without the walker. Still, they took more steps and initiated more locomotor bouts in the walker condition; losing access to crawling did not outweigh the increase in upright stepping activity. Even though suspension walkers boosted locomotor activity for infants of all locomotor statuses, the walker provided a larger advantage for young infants who could not yet crawl or cruise compared to infants who could. Thus, suspension walkers may be particularly effective at facilitating locomotor opportunities before infants acquire independent locomotor skills.

Caregivers played an active role in facilitating locomotor behavior by moving the walker: Caregiver-initiated movement accounted for the majority of walker movement and infant stepping rates were much higher when the walker was moving compared to stationary. Stepping was likely higher when the walker was moving because the moving floor prompts stepping as a response. Indeed, even 3-day-old typically developing infants step more when held over a moving treadmill belt compared to a stationary treadmill belt (Siekerman et al., 2015). Moreover, on treadmills, infants step more when provided with optic flow consistent with self-movement (Barbu-Roth et al., 2009; Moerchen & Saeed, 2012) and when more aroused (Thelen et al., 1982). Moving in the walker provides paired optic flow and may be more arousing and engaging than remaining stationary, thus further increasing stepping response beyond just the prompting provided by the moving floor. Indeed, in our own prior work, we found that infants stepped more when pushed in the walker compared to held over a moving treadmill belt (Hospodar et al., 2026), suggesting that movement through space is indeed important for facilitating infant stepping.

Despite the prevalence of caregiver-initiated movement, we found that over half of the infants at every age self-propelled the walker. Critically, these results of self-propulsion are higher than what we found when we explicitly tested self-propulsion in prior work. One major difference is that in the explicit test of self-propulsion, we locked the wheels of the walker so that the wheels could not move backwards, but the wheels were unlocked for free play. Thus, moving backwards provides another option for self-propulsion (in the current study, infants moved backwards for 45% of their infant-initiated bouts, forward for 37%, and in both directions for 18%). Regardless of direction, our results suggest infants can self-initiate movement with a walker as young as 10 months of age and before they are able to crawl or cruise. Being able to move on their own terms—even backwards—may encourage infants to continue trying to move, thereby gaining practice.

### Limitations and future directions

Due to time restrictions, we could only collect approximately six minutes of free play data per condition. Data were also collected in a standardized playroom, which may be different from real-world settings. Thus, we do not yet know how long infants can tolerate the walker before they become fussy or fatigued, or how caregivers would engage with the walker on longer timescales or in varied environments. Critically, our results represent infants’ immediate responses to the suspension walker without practice. With continued use, we would expect that infants would improve at using the walker for self-initiated locomotion. In future work, we plan to examine changes in self-initiated walker use and locomotor behavior with repeated practice.

## Conclusions

Our results provide preliminary evidence for the provision of overground support to pre-walking infants with DS: Suspension walkers robustly increased infants’ immediate locomotor behavior, even without prolonged practice. Caregivers played an active role in facilitating infants’ activity by moving the walker, and walker movement encouraged infant stepping. Thus, our results suggest that overground walkers can support children’s everyday activity in their natural environment. Future work will directly examine the potential of overground suspension walkers for home-based intervention.

## Data Availability

All data produced in the present study will be available on Databrary (https://databrary.org) upon publication.

