## Supplement for "Immediate effects of suspension walker support on locomotor behavior of pre-walking infants with Down syndrome"

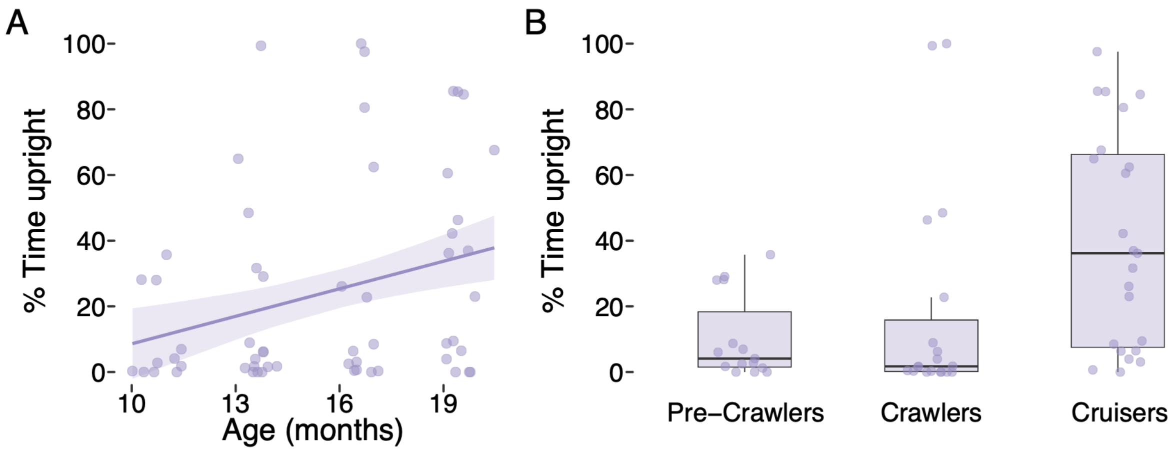


Supplemental Figure 1. Percent time upright in the no-walker condition by (A) age and (B) locomotor status. (A) The banded line represents the model estimation with 95% confidence interval. Circular points represent individual observations across visits from each infant. (B) The height of each box plot extends to the interquartile range and the horizontal black line represents the median for each group; whiskers extend to 1.5 times the interquartile range. Circular points represent individual observations across visits from each infant.
